# Healthcare Workers’ Knowledge, Attitudes, and Practices Towards Electronic Medical Records in Ogun State, Nigeria

**DOI:** 10.1101/2025.08.20.25334058

**Authors:** Akingbade Rapheal Anuoluwapo

## Abstract

Electronic Medical Records (EMRs) are critical for improving healthcare delivery, yet their adoption in low-resource settings faces multiple challenges. In 2023, Ogun State, Nigeria, introduced EMRs in one tertiary and five secondary hospitals. This study assessed healthcare workers’ knowledge, attitudes, and practices (KAP) regarding EMRs to identify barriers and enablers for effective implementation. A cross-sectional descriptive study was conducted among 330 healthcare professionals, including doctors, nurses, pharmacists, laboratory scientists, and health records officers. Data were collected using structured questionnaires and key informant interviews and analyzed using descriptive statistics, chi-square tests, logistic regression, and thematic content analysis. Findings revealed moderate knowledge of EMR functions (65%), positive attitudes toward adoption (76%), but low consistent practice, with only 28% fully relying on EMRs due to infrastructural challenges such as power outages (85%), poor internet connectivity (70%), and inadequate training (65%). Facilitators included perceived efficiency, improved patient safety, and government support. The study concludes that while healthcare workers in Ogun State are willing to adopt EMRs, infrastructural and capacity-related barriers hinder full utilization. Targeted interventions focusing on ICT infrastructure, training, and policy support are recommended to enhance adoption and sustainability.

## Introduction

Electronic Medical Records (EMRs) are digital versions of patient charts that enhance documentation, clinical decision-making, and patient care continuity (WHO, 2020). EMRs are integral to health system efficiency, patient safety, and evidence-based practice. Globally, high-income countries such as the USA and UK have achieved high EMR adoption through policy mandates and financial incentives (Adler-Milstein & Jha, 2017). In contrast, low- and middle-income countries (LMICs) often face infrastructural, financial, and workforce challenges (Oleribe et al., 2019).

In Nigeria, despite policy frameworks like the National Health ICT Strategic Framework (2016–2020), EMR implementation remains limited. Ogun State initiated EMR deployment in 2023 across six hospitals, representing a significant digital health investment. However, anecdotal reports suggest healthcare workers’ use remains inconsistent due to infrastructural deficits, limited training, and varying levels of digital literacy. Assessing healthcare workers’ knowledge, attitudes, and practices (KAP) provides insight into behavioral and systemic factors influencing EMR adoption, informing policy and capacity-building strategies.

### Study Objectives

- Assess knowledge of EMRs among healthcare workers.
- Examine attitudes toward EMR adoption.
- Evaluate current practices of EMR use.
- Identify barriers and facilitators influencing EMR utilization.

## Methods

A cross-sectional descriptive study was conducted in six Ogun State hospitals, including Olabisi Onabanjo University Teaching Hospital (OOUTH) and five secondary facilities. The population comprised 1,200 healthcare workers across various cadres. Using Yamane’s formula (1967) with a 5% margin of error, a sample size of 300 was calculated, increased to 330 to account for non-responses.

### Sampling & Data Collection

Stratified random sampling ensured proportional representation across professional cadres. Data were collected via structured questionnaires covering socio-demographics, knowledge, attitudes, practices, and perceived barriers/facilitators. Key informant interviews (KIIs) with hospital administrators explored system implementation experiences.

### Data Analysis

Quantitative data were analyzed with SPSS (v25) using descriptive statistics, chi-square tests, and logistic regression to determine predictors of positive attitudes and EMR use. Qualitative data were coded and thematically analyzed using NVivo.

### Ethical Considerations

Approval was obtained from the Ogun State Hospital Management Board Ethical Review Committee. Participation was voluntary, with informed consent and confidentiality assured.

## Results

### Socio-Demographic Profile

Among 330 respondents, 52% were female, and 48% male; 36% were nurses, 24% doctors, 18% health records officers, 12% laboratory scientists, and 10% pharmacists. Most respondents (38%) were aged 30–39 years, with 32% having 6–10 years of experience.

### Knowledge

- 82% were aware of the EMR rollout.
- 65% demonstrated knowledge of core EMR functions (patient registration, clinical documentation, prescription management).
- Only 32% understood advanced functionalities such as decision support and interoperability.

### Attitudes

- 76% perceived EMRs as improving patient care and reducing errors.
- 70% agreed EMRs save time, although 55% worried about system downtime and power outages.

### Practices

- 60% reported hybrid use (paper + EMR).
- Only 28% consistently relied on EMRs.
- Real-time data entry was reported by 40% of respondents.

### Barriers & Facilitators

- Key barriers: power outages (85%), poor internet connectivity (70%), inadequate training (65%), workload pressures (55%), and resistance among senior staff (30%).
- Facilitators: perceived efficiency, improved patient safety, government support, and time savings.

## Discussion

Healthcare workers in Ogun State demonstrated moderate knowledge, positive attitudes, but low EMR utilization. Knowledge gaps in advanced EMR functionalities may hinder optimal use. Positive attitudes suggest willingness to adopt, consistent with the Technology Acceptance Model (Davis, 1989).

Practice gaps are largely due to infrastructural limitations, confirming trends reported in LMICs such as Kenya, Ghana, and Ethiopia (Were et al., 2010; Osei-Tutu et al., 2018; Tilahun & Fritz, 2015). Barriers identified align with previous findings, emphasizing that policy and system support are critical to successful EMR adoption. Facilitators highlight opportunities for leveraging government investment and staff motivation to improve uptake.

### Implications

- ICT infrastructure and reliable power are prerequisites for sustained EMR use.
- Continuous training and dedicated IT support can bridge knowledge-practice gaps.
- Institutional policy support ensures system integration and long-term sustainability.

## Conclusion

Healthcare workers in Ogun State show readiness to embrace EMRs, yet systemic and infrastructural challenges limit consistent use. Strengthening ICT infrastructure, implementing ongoing training, and enhancing policy support are essential to realizing EMRs’ potential for improved healthcare delivery.

## Recommendations

1. **Government & Policymakers**: Invest in reliable power, internet, and hardware; integrate EMRs into broader health information systems.
2. **Hospital Management**: Institutionalize EMR training, appoint IT support staff, and phase out paper-based systems gradually.
3. **Healthcare Workers**: Engage in continuous learning, provide feedback on system usability, and champion EMR adoption within clinical teams.
4. **Researchers**: Conduct longitudinal studies to monitor KAP changes, explore patient perspectives, and assess cost-effectiven

## Data Availability

All data produced in the present study are available upon reasonable request to the authors

## References

Adler-Milstein, J., & Jha, A. K. (2017). HITECH Act drove large gains in hospital electronic health record adoption. Health Affairs, 36(8), 1416– 1422.10.1377/hlthaff.2016.1651

Ajami, S., & Bagheri-Tadi, T. (2013). Barriers for adopting electronic health records (EHRs) by physicians. Acta Informatica Medica, 21(2), 129– 134. 10.5455/aim.2013.21.129-134

Adeleke, I. T., Olowu, A. A., & Adedokun, O. T. (2012). Assessment of EMR implementation in Nigerian tertiary hospitals. Journal of Health Informatics in Developing Countries, 6(2), 22–31.

Davis, F. D. (1989). Perceived usefulness, perceived ease of use, and user acceptance of information technology. MIS Quarterly, 13(3), 319–340. 10.2307/249008

Oleribe, O. O., et al. (2019). Electronic health record adoption in Sub-Saharan Africa: Challenges and opportunities. Journal of Public Health in Africa, 10(2), 1023.

Osei-Tutu, E., et al. (2018). EMR adoption in Ghana: Barriers and facilitators. BMC Medical Informatics and Decision Making, 18(1), 100.

Tilahun, B., & Fritz, F. (2015). Factors influencing the implementation of EMR systems in Ethiopia. BMC Medical Informatics and Decision Making, 15, 39.

Were, M., et al. (2010). EMR adoption in Kenyan HIV clinics: Knowledge, attitude, practice survey. International Journal of Medical Informatics, 79(12), 873–884.

World Health Organization. (2020). Global strategy on digital health 2020–2025. Geneva: WHO.

